# Healthcare services access, use, and barriers among migrants in Europe: a systematic review

**DOI:** 10.1101/2022.02.24.22271449

**Authors:** Petros Galanis, Koureas Spyros, Olga Siskou, Olympia Konstantakopoulou, Georgios Angelopoulos, Daphne Kaitelidou

**Author notes:** **Corresponding author:** Petros Galanis, Assistant Professor, Clinical Epidemiology Laboratory, Faculty of Nursing, National and Kapodistrian University of Athens, Greece, 123 Papadiamantopoulou street, GR-11527, Athens, Greece. **Funding:** National and Kapodistrian University of Athens and “Stavros Niarchos Foundation”.

## Abstract

**Background:** The issue of migrants health and access to health services is dynamic and complex posing a challenge to health systems worldwide.

**Aim:** To investigate migrants’ access to health services in European countries, the use of health services by migrants and the barriers encountered by migrants in the use of health services.

**Material and methods:** The search was conducted in January 2022 in five databases; PubMed, Medline, Web of science, Scopus and Cinahl. We used the following keywords: migrants, immigrants, use, access, utilization, healthcare services, services, needs, health, difficulties, barriers. The inclusion criteria were the following: (a) the studies investigated the access of migrants to health services, the use of health services by migrants and the barriers encountered by migrants in using health services. (b) migrants self-assessed access, use and barriers. (c) studies were conducted in European countries. (d) studies included adult migrants. (e) the language of articles was English.

**Results:** Sixty-five studies were met our inclusion criteria. among studies, 89.2% were quantitative and 11.8% were qualitative. All quantitative studies were cross-sectional. for data collection, 58.5% of studies used questionnaires and 30.8% used historical files. Also, personal interviews were performed in 9.2% of studies and focus groups in 1.5% of studies. in our studies, 73.8% of natives stated that they had better access to health services and used health services better than migrants, while 26.2% found that migrants stated that they had better access to health services and used health services better. The most common barriers were the following: inability to understand the language and communicate, lack of insurance, lack of information and knowledge, lack of family support, low educational level, short duration of stay in the country of migration, low income, lack of a family doctor and high costs.

**Conclusions:** Migrants face several barriers both in accessing and using health services in Europe. Intensive efforts are needed to increase migrants’ knowledge, implement culturally sensitive interventions in migrant communities and better inform healthcare professionals so that they can approach migrants more effectively.

## Introduction

The high mobility of populations, the economic crisis and the significant increase in unemployment have defined the last decades, with the economic crisis affecting vulnerable groups, such as migrants. Thus, a particular aspect of health inequalities has emerged, which mainly concerns the barriers and difficulties encountered by migrants in accessing healthcare services, which has a direct impact on the quality of the services provided. The Americas is the continent with the largest number of migrants, with the number increasing from 47 million in 2000 to 60 million in 2012 (Loue & Sajatovic, 2012). The situation is similar in Europe, with migrants making up around 9% of the total population (Loue & Sajatovic, 2012). In fact, in Europe, 7-13% of migrants do not have legal residence documents (Karl-Trummer et al., 2010).

Access to high quality health services is an extremely important issue for migrants, as they face unequal opportunities for treatment interventions (The Lancet Public Health, 2018). The growing migrant population is also reflected in the increasing need for health services. For this reason, a corresponding adjustment of health policies at international level is required to better provide health care for migrants (Macpherson et al., 2007). Indeed, in recent years the issue of migrants’ health has expanded from providing health care to patients to health promotion and prevention. The World Health Organization plays an essential role in better addressing health issues related to migrants (World Health Organization, 2008).

The issue of migrants health and access to health services is dynamic and complex, involving different stages of migration such as the pre-migration period, the first years of migration and migrants after several years of residence in the host country (Lassetter & Callister, 2009; Ullmann et al., 2011). Furthermore, this issue is also related to various social determinants that are not only related to the characteristics of migrants, e.g. gender, cultural diversity, experiences, etc., but also to the cultural environment of the host countries, e.g. the health care system, living conditions, moral, religious and cultural values, etc. (Malmusi et al., 2010; Spallek et al., 2011).

Migrants and their children are less often insured and use health services less often than natives (Abe-Kim et al., 2007; Alegría et al., 2006; Callahan et al., 2006; Cunningham, 2006; Guendelman et al., 2001; Huang et al., 2006; Jackson et al., 2007; Javier et al., 2007; Lasser et al., 2006; Yu et al., 2006). In addition, the cost of health care for migrants is lower, with the exception of the cost of emergency care for migrant children (Derose et al., 2009).

The aim of this systematic literature review was to investigate migrants’ access to health services, the use of health services by migrants and the barriers encountered by migrants in the use of health services.

## Material and Methods

The search was conducted in January 2022 in five databases; PubMed, Medline, Web of Science, Scopus and Cinahl. We used the following key-words: migrants, immigrants, use, access, utilization, healthcare services, services, needs, health, difficulties, barriers.

The inclusion criteria were the following: (a) the studies investigated the access of migrants to health services, the use of health services by migrants and the barriers encountered by migrants in using health services. (b) migrants self-assessed access, use and barriers. (c) studies were conducted in European countries. (d) studies included adult migrants. (e) the language of articles was English

Studies that included in the systematic literature review are presented in Table 1, while the flowchart is presented in Figure 1.

**Table 1.**
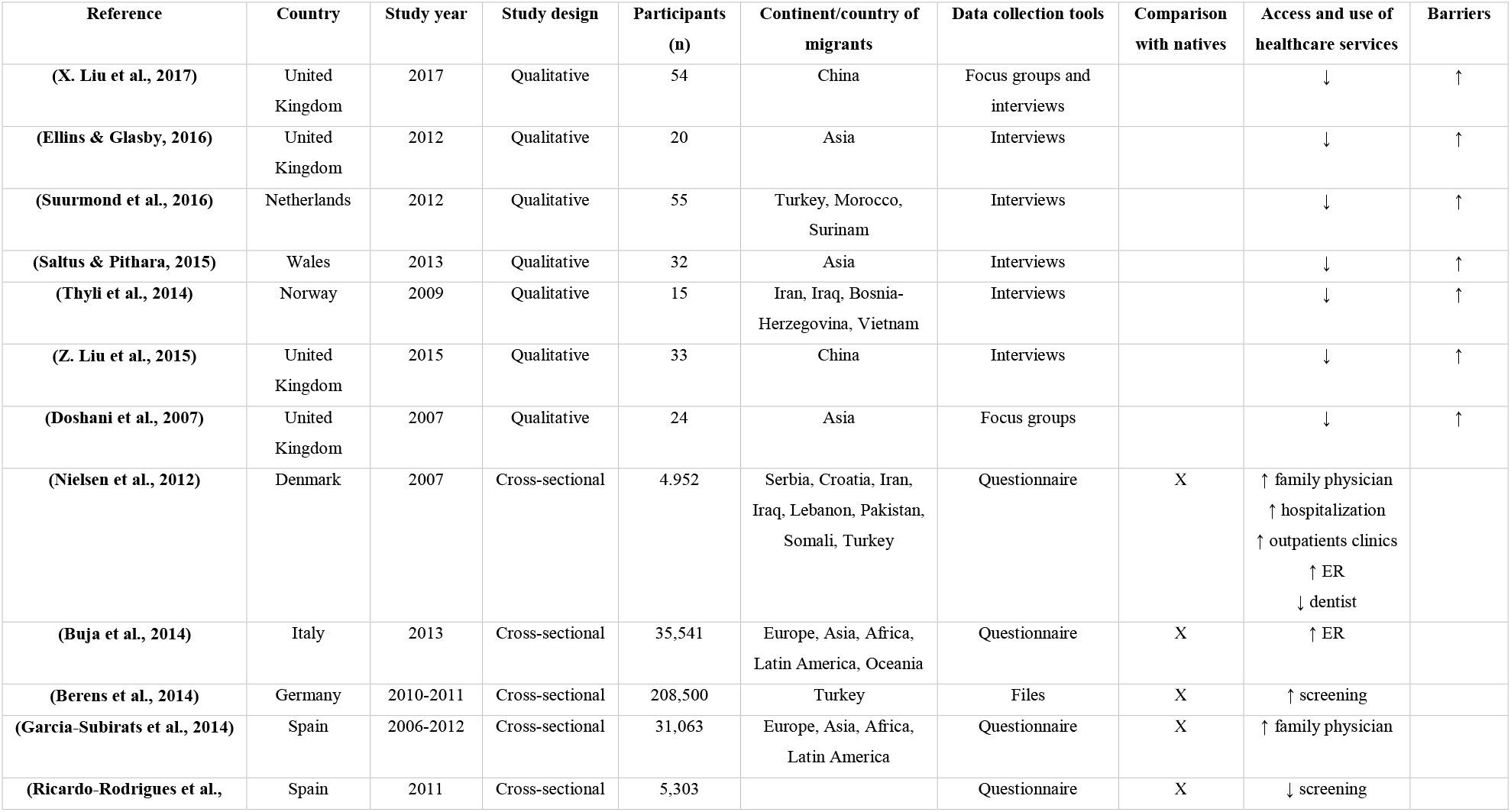
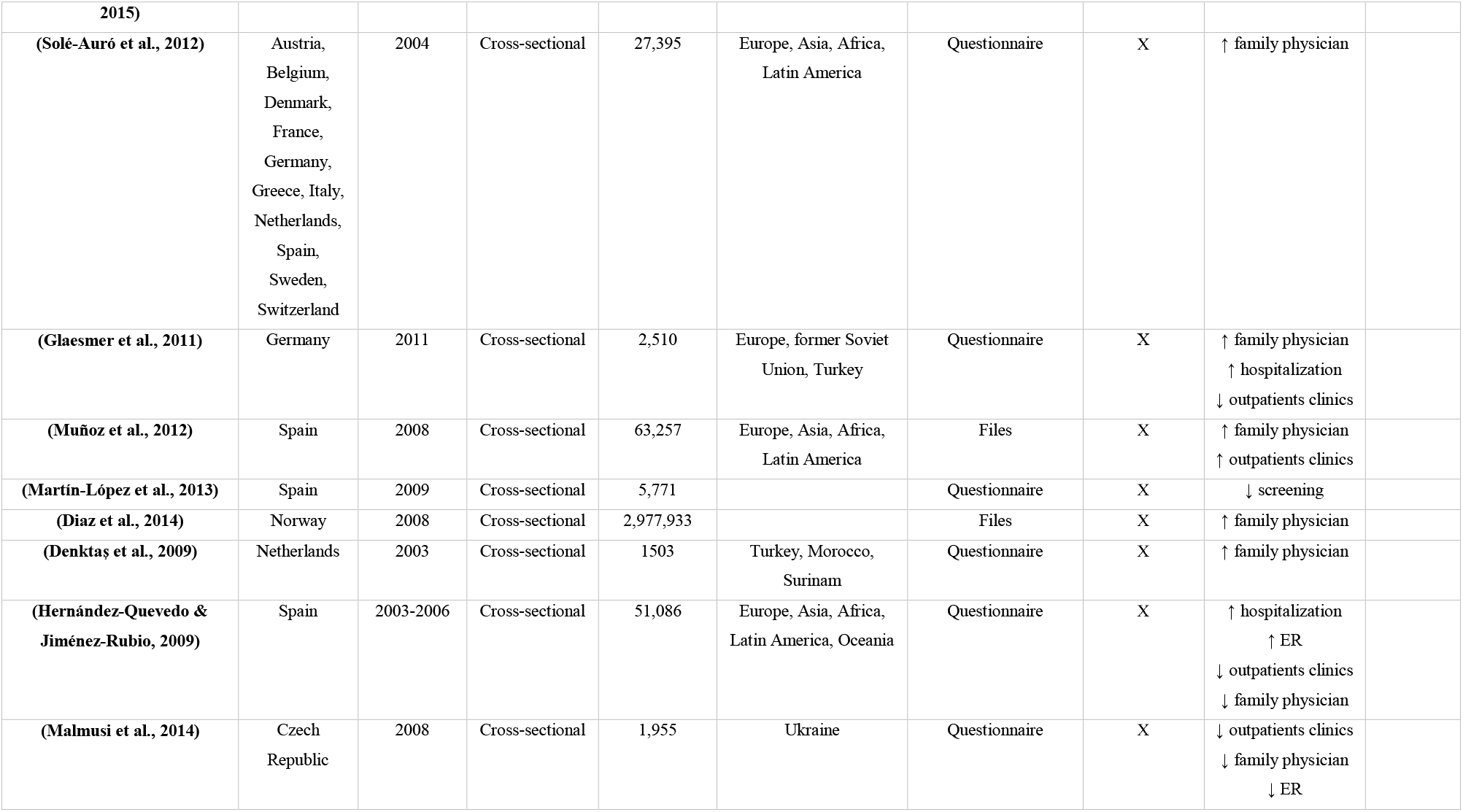
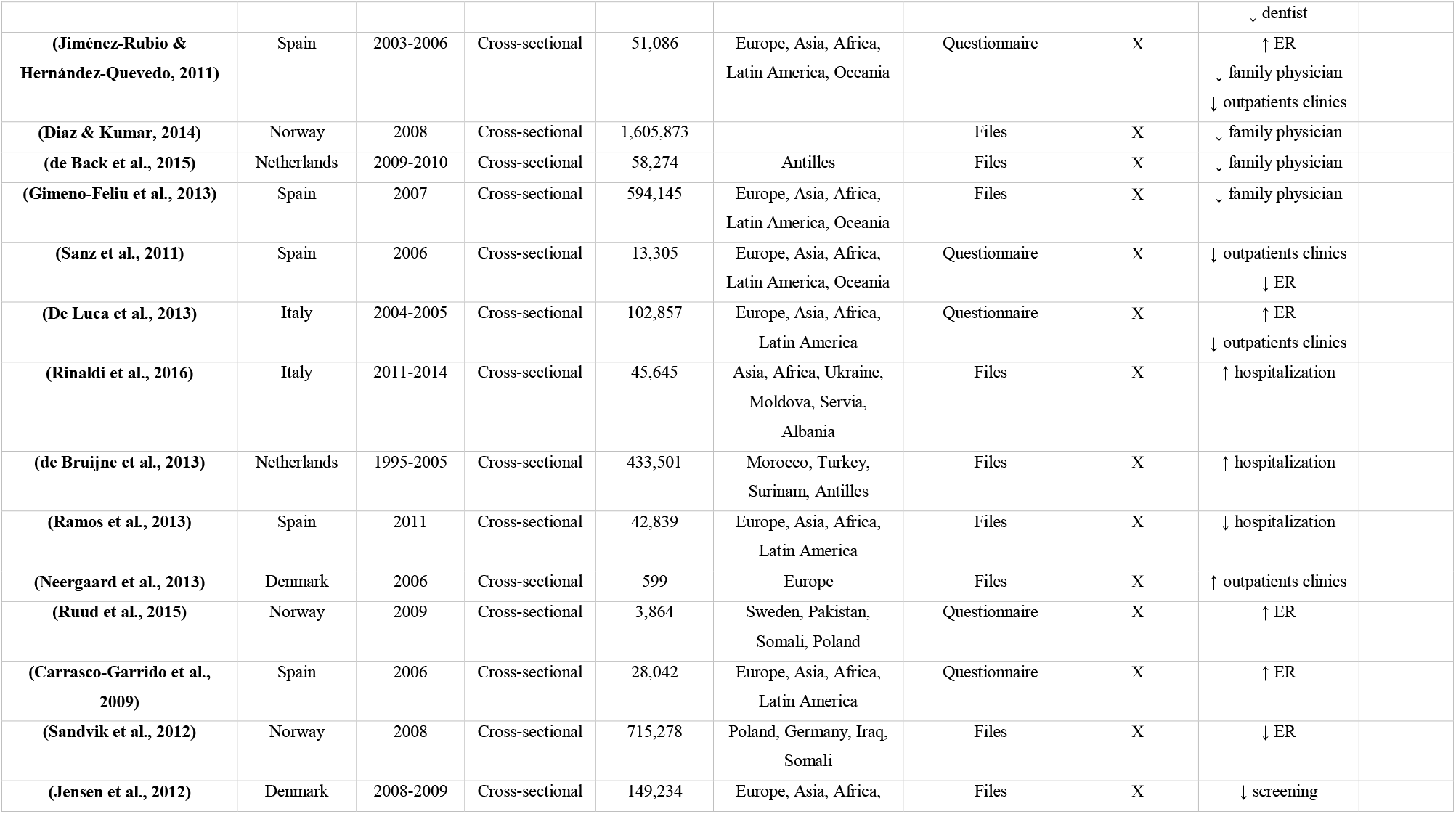
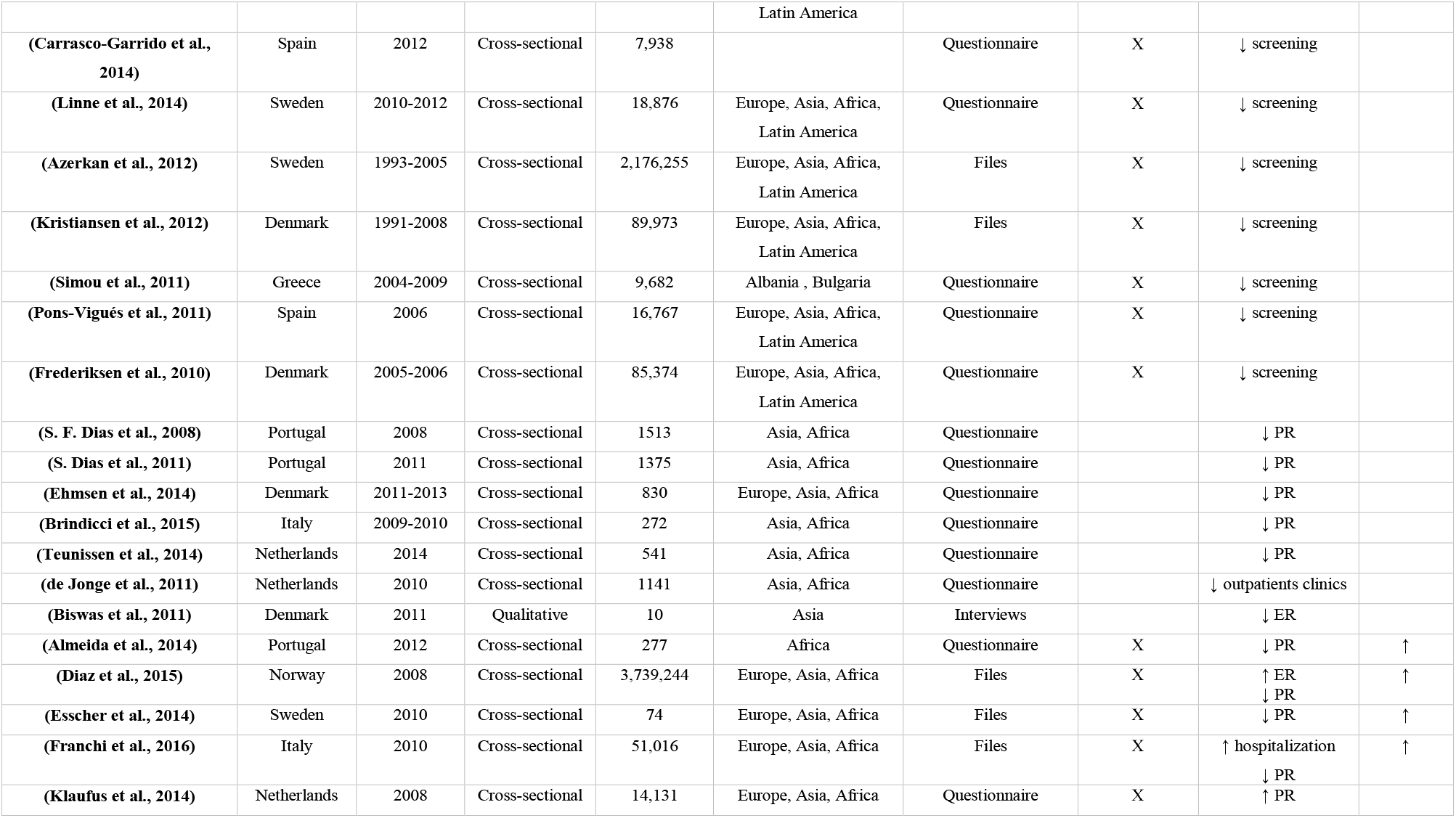
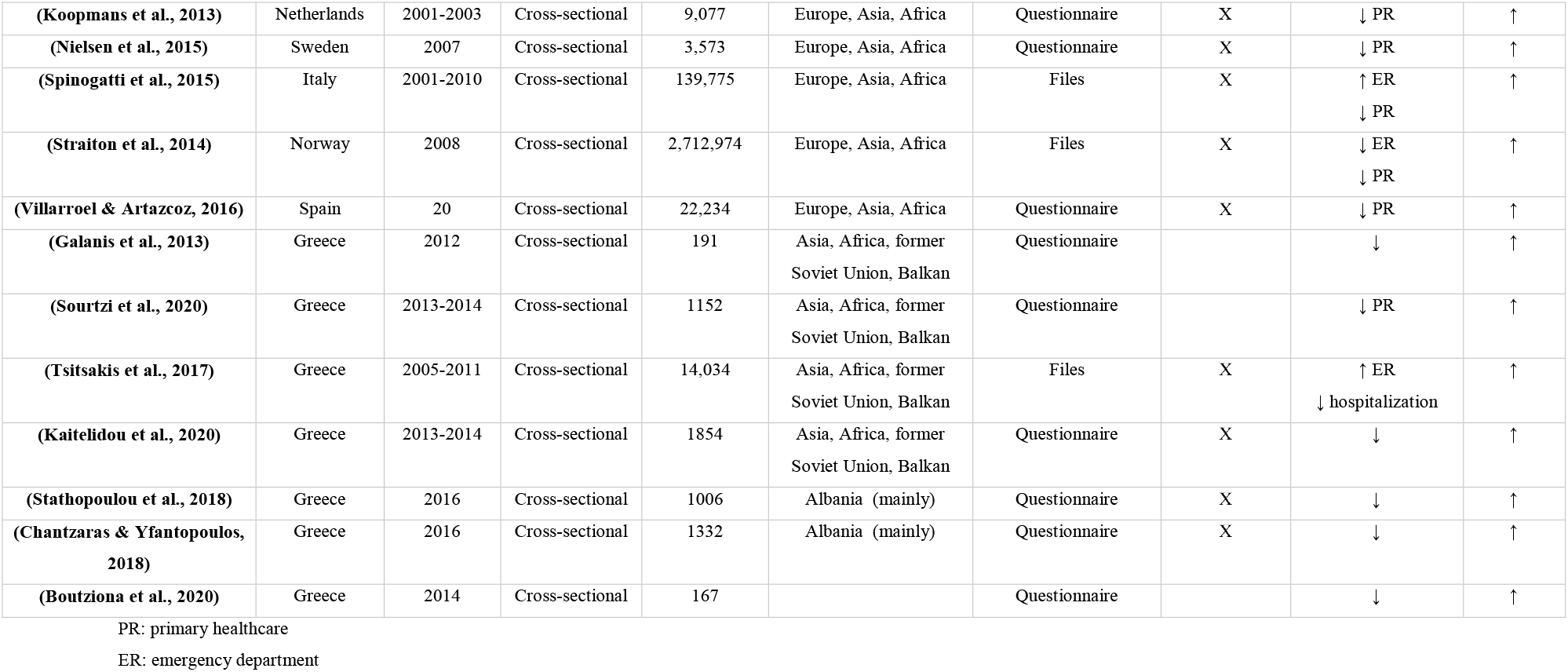
Studies on migrants’ access to health services, use of health services and the relative barriers.

**Figure 1.**
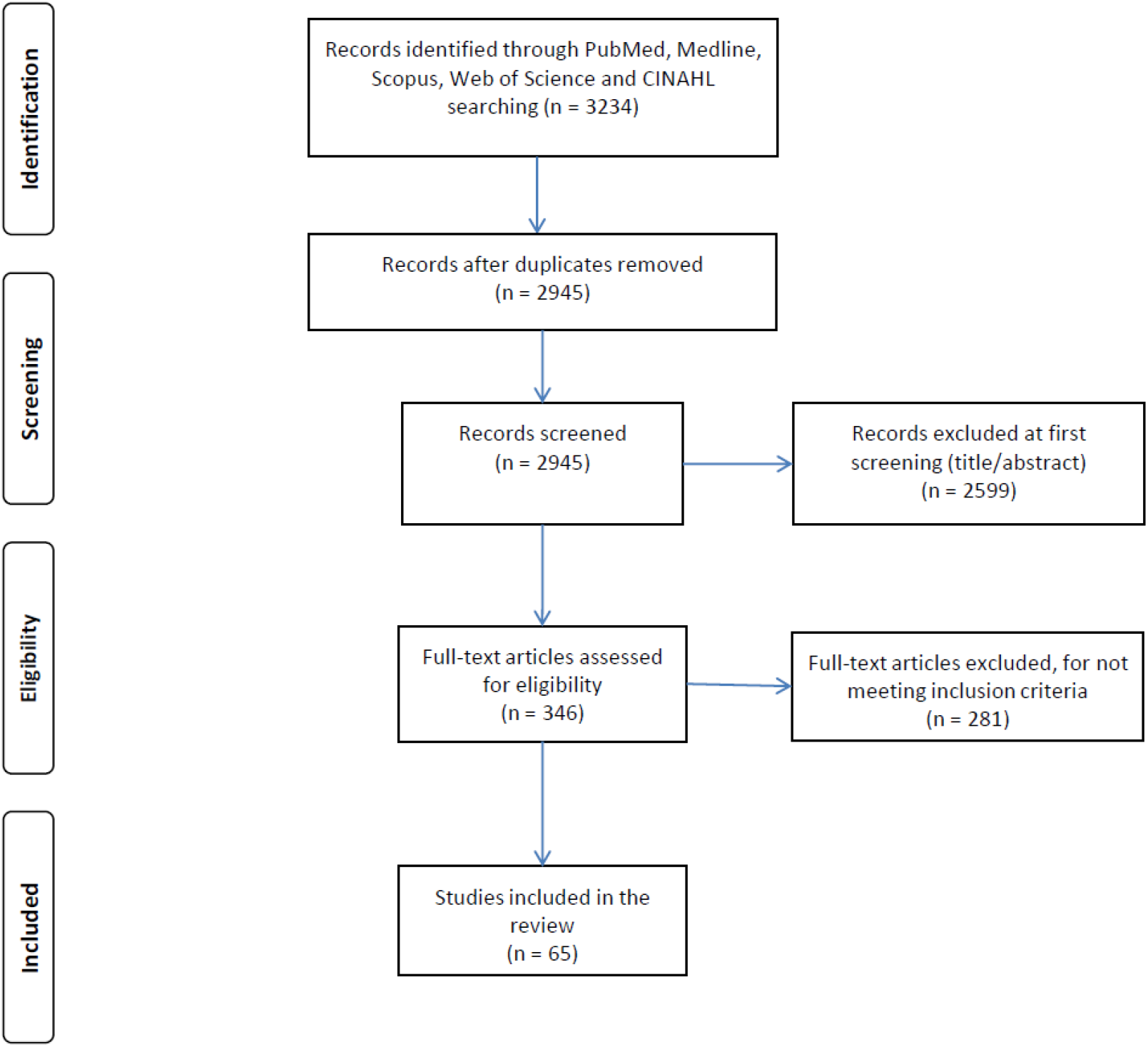
Flowchart of the systematic review.

## Results

Sixty-five studies were met our inclusion criteria. Countries that studies were conducted in our review were Spain, Italy, United Kingdom, Greece, Austria, Belgium, France, Netherlands, Wales, Portugal, Germany, Denmark, Switzerland, Norway, Sweden and Czech Republic.

Among studies, 89.2% were quantitative and 11.8% were qualitative. All quantitative studies were cross-sectional. In qualitative studies, the minimum sample size was 10 and the maximum number was 55, while in the quantitative studies, the minimum sample size was 74 and the maximum number was 3,739,244.

For data collection, 58.5% of studies used questionnaires and 30.8% used historical files. Also, personal interviews were performed in 9.2% of studies and focus groups in 1.5% of studies.

Natives were also included in 73.8% of studies allowing comparisons with migrants. In these studies, 73.8% of natives stated that they had better access to health services and used health services better than migrants, while 26.2% found that migrants stated that they had better access to health services and used health services better.

Among studies, 18.5% investigated migrants’ access to health services in general, while 81.5% investigated access to specific health services, such as primary health care services, general practitioner, emergency department, hospitalization, etc.

The barriers faced by migrants in accessing and using health services are shown in Table 2. The most common barriers were the following: inability to understand the language and communicate, lack of insurance, lack of information and knowledge, lack of family support, low educational level, short duration of stay in the country of migration, low income, lack of a family doctor and high costs.

**Table 2.** Detailed description of the barriers encountered by migrants in accessing and using healthcare services.

| Reference | Language and communication difficulties | Lack of information and knowledge | Family support failure | Cultural and religion issues | Fear for pain | Difficulties to understand physicians' guidelines |
| --- | --- | --- | --- | --- | --- | --- |
| (X. Liu et al., 2017) | X | X | X | X |  |  |
| (Galanis et al., 2013) | X |  |  |  |  |  |
| (Boutziona et al., 2020) | X |  |  | X |  |  |
| (Koopmans et al., 2013) | X |  | X |  |  |  |
| (Nielsen et al., 2015) | X |  | X |  |  |  |
| (Spinogatti et al., 2015) |  |  | X |  |  |  |
| (Straiton et al., 2014) |  |  | X |  |  |  |
| (Villarroel & Artazcoz, 2016) |  |  | X |  |  |  |
| (Beiser & Hou, 2014) | X |  | X |  |  |  |
| (Diaz et al., 2015) |  |  | X |  |  |  |
| (Durbin et al., 2014) | X |  |  |  |  |  |
| (Esscher et al., 2014) | X |  |  |  |  |  |
| (Klaufus et al., 2014) |  |  | X |  |  |  |
| (Almeida et al., 2014) |  |  | X |  |  |  |
| (Ho et al., 2005) |  |  | X |  |  |  |
| (Ellins & Glasby, 2016) | X | X |  |  |  |  |
| (Suurmond et al., 2016) | X | X | X |  |  |  |
| (Thyli et al., 2014) | X |  | X |  | X |  |
| (Z. Liu et al., 2015) | X |  |  |  |  | X |
| (Doshani et al., 2007) |  | X |  |  |  |  |
| (Garcia-Subirats et al., 2014) |  |  |  |  | X |  |
| (Denktaş et al., 2009) | X |  |  |  |  |  |

**Table 2 (continued).** Detailed description of the barriers encountered by migrants in accessing and using healthcare services.
| Παραπομπή | Females shame and modesty | Long waiting hour | Lack of time | High cost | Lack of insurance | Emotional barriers | Functional disability |
| --- | --- | --- | --- | --- | --- | --- | --- |
| (Galanis et al., 2013) |  | X |  | X |  | X |  |
| (Boutziona et al., 2020) |  |  |  | X | X |  |  |
| (Almeida et al., 2014) |  |  |  |  | X |  |  |
| (Doshani et al., 2007) | X |  |  |  | X |  |  |
| (Garcia-Subirats et al., 2014) |  | X | X | X | X |  |  |
| (Denktaş et al., 2009) |  |  |  |  | X |  |  |
| (X. Liu et al., 2017) |  | X |  |  | X |  |  |
| (Sourtzi et al., 2020) | X |  |  |  | X |  |  |
| (Boutziona et al., 2020) | X |  |  |  | X |  |  |
| (Esscher et al., 2014) | X |  |  |  | X |  |  |
| (Klaufus et al., 2014) | X |  |  |  |  |  | X |
| (Nielsen et al., 2015) | X |  |  |  |  |  | X |
| (Spinogatti et al., 2015) | X |  |  |  |  |  |  |
| (Villarroel & Artazcoz, 2016) | X |  |  |  |  |  |  |
| (Jiménez-Rubio & Hernández-Quevedo, 2011) | X |  |  |  |  |  |  |
| (Garcia-Subirats et al., 2014) | X |  |  |  |  |  |  |
| (Suurmond et al., 2016) |  |  | X |  |  |  |  |
| (Denktaş et al., 2009) |  |  | X |  |  |  |  |

**Table 2 (continued).** Detailed description of the barriers encountered by migrants in accessing and using healthcare services.
| Παραπομπή | Short length of stay | Low income | Stay in rural areas | Low educational level | Limited social integration | Lack of trust | Self-esteem of good health status |
| --- | --- | --- | --- | --- | --- | --- | --- |
| (Saltus & Pithara, 2015) |  |  |  |  | X |  |  |
| (Spinogatti et al., 2015) |  |  |  | X |  |  |  |
| (Straiton et al., 2014) | X | X |  |  |  |  |  |
| (Villarroel & Artazcoz, 2016) |  |  |  | X |  |  |  |
| (Sourtzi et al., 2020) |  | X |  |  |  |  |  |
| (Beiser & Hou, 2014) | X |  |  |  |  |  |  |
| (Diaz et al., 2015) |  | X | X | X |  |  |  |
| (Durbin et al., 2014) |  | X |  | X |  |  |  |
| (Esscher et al., 2014) |  | X |  | X |  |  |  |
| (Klaufus et al., 2014) |  | X |  | X | X |  |  |
| (Koopmans et al., 2013) |  |  |  | X |  |  |  |
| (Nielsen et al., 2015) | X | X |  |  |  |  |  |
| (Almeida et al., 2014) |  | X |  | X |  |  |  |
| (Ricardo-Rodrigues et al., 2015) |  |  | X | X |  |  |  |
| (Solé-Auró et al., 2012) |  |  |  |  |  |  |  |
| (Diaz et al., 2014) | X | X |  |  |  |  |  |
| (Jiménez-Rubio & Hernández-Quevedo, 2011) |  | X |  |  |  |  |  |
| (Diaz & Kumar, 2014) | X |  |  |  |  |  |  |
| (Kristiansen et al., 2012) |  |  |  | X |  |  |  |
| (Pons-Vigués et al., 2011) |  | X |  |  |  |  |  |
| (Jiménez-Rubio & Hernández-Quevedo, 2011) |  |  |  |  |  |  | X |
| (Garcia-Subirats et al., 2014) |  |  |  |  |  |  | X |
| (Thyli et al., 2014) |  |  |  |  |  | X |  |

The barriers that migrants face in accessing and using health services can be summarized in the following five broad categories: social and economic barriers, health system-related barriers, cultural barriers, knowledge-related barriers and personal barriers.

Social barriers included low educational attainment, lack of insurance, high costs, low income, frequent travel, limited social integration and inability to understand language and communication.

Barriers related to the health system included lack of a family doctor, limited understanding of doctors’ instructions, long waiting time, difficulties in access, lack of translators and living in rural areas.

Cultural barriers included stigma, shame and modesty on the part of females and religious beliefs.

Knowledge-related barriers included lack of knowledge about how to access and navigate the health system, lack of knowledge about screening, perception that screening is unnecessary and self-perception of good health status.

Personal barriers include lack of family support, short length of stay in the country of migration, fear, lack of time, anxiety, lack of confidence and functional incapacity.

## Discussion

According to our review, migrants face several barriers both in accessing and using health services.

Firstly, there are significant social and economic barriers that limit migrants’ access to health services, as well as their use. More specifically, low social and economic level, as well as living in areas characterized by low income, is a determining factor that reduces the use of health services by migrants, especially those services related to screening, such as in the case of mammography (Ahmad et al., 2012; Donnelly et al., 2009). In many cases, migrants with low social and economic status are paid an hourly rate, resulting in the use of health services leading to lost time and thus reduced income (Donnelly et al., 2009; Meana et al., 2001). This is supported by the fact that moving to health services requires some costs. A typical example is women with a low level of education and limited social integration, who make very little use of screening services (Donnelly et al., 2009; Meana et al., 2001). It is noted that low educational and economic level also leads to a lack of knowledge and understanding which further limits the use of health services by migrants. It is clear that the high cost of some services acts as a deterrent to their use by migrants, as they are unable to cover it (Lobb et al., 2013).

Inability to understand the language is an extremely important barrier faced by migrants, as it leads to inability to communicate, frustration and ultimately abandonment of efforts to use health services (Ahmad et al., 2012; Sun et al., 2010; Todd et al., 2011; Woloshin et al., 1997). Language difficulties reduce the ability of migrants to communicate with healthcare staff and understand their instructions. In addition, the ability of migrants to inform themselves and increase their knowledge, such as the ability to browse websites related to health services or screening, is also significantly limited. Unfortunately, an important obstacle related to the health care system also contributes to this, which is the lack of translators who could improve the quality of communication between migrants and medical staff.

The lack of a family doctor is another important barrier related to the health system, especially in the case of screening, such as in the case of mammography (Ahmad et al., 2012; Meana et al., 2001; Steven et al., 2004; Sun et al., 2010; Todd et al., 2011; Vahabi et al., 2016). Moreover, in some cases, the increased workload of doctors does not allow them to spend more time with migrants to explain more to them and answer their questions and queries especially in the case of screening. In fact, the lack of translators further exacerbates this problem, as doctors are unable to communicate with migrants, with the result that doctors’ instructions and recommendations are not understood by migrants.

Particular reference needs to be made to the cultural barriers that migrants face in using health services. A typical example is the case of migrant women with particular religious beliefs. In this case, the existence of male doctors is a particular inhibiting factor for the use of health services related to sensitive issues, such as mammography (Ahmad et al., 2012; Steven et al., 2004; Todd et al., 2011; Vahabi et al., 2016). Some religions recognize women’s bodies as ‘sacred’ and ‘private’, with the result that women may feel shame and guilt when a male doctor has to look at their bodies in the case of, for example, mammography. Many women, moreover, believe that the occurrence of a disease is God’s will or the result of fate, so they do not use screening methods as they consider them unnecessary. In even more extreme situations, some societies consider breast cancer to be the result of immorality, so they view mammography negatively. The problem is further exacerbated by the fact that in some cases medical staff avoid discussing sensitive issues with migrants, in order to respect their cultural sensitivity and avoid possible friction and misunderstandings. This problem is extremely important, as in many cases the recommendation from health professionals is the most important motivation for migrants to undergo screening (Hanson et al., 2009). It is a fact that in most countries not much attention is paid to the cultural and religious specificities of migrants, which, especially in the case of the use of health services, are a decisive factor (Lobb et al., 2013).

Lack of knowledge is a key barrier to the use of health services by migrants, particularly in the case of screening. For example, migrant women are largely unaware of the risk factors for breast cancer (Ahmad et al., 2012; Meana et al., 2001; Vahabi et al., 2016). It is common, moreover, for migrant women to seek medical help only if they develop symptoms and signs, as health systems in their countries mainly emphasize treatment rather than prevention (Lobb et al., 2013). Similarly, many migrant women have limited knowledge about the benefits, side effects and the internetAsia of mammography and other screening methods. Thus, the lack of knowledge leads migrants to not use screening services as they consider them unnecessary (Hyman et al., 2001). The problem is magnified in the case of migrants who self-assess their health status as good, so that they consider that screening is not necessary (Garcia-Subirats et al., 2014; Jiménez-Rubio & Hernández-Quevedo, 2011). Moreover, the lack of knowledge about how to access and navigate the health system creates even more problems, as migrants do not even know how to access the different services. It should be noted that the lack of knowledge is often combined with a lack of understanding of the language and a lack of translators in health services (Lobb et al., 2013).

Many migrants do not undergo screening because they are concerned about the consequences of being diagnosed with cancer, such as stress, access to necessary services, social isolation, poor prognosis, etc. (Ahmad et al., 2012; Vahabi et al., 2016). In addition, due to lack of knowledge, migrants are also concerned about the adverse effects of different tests, such as exposure to radiation during a mammogram, adverse effects of a therapeutic intervention (e.g. surgical removal of a tumor), etc. (Ahmad et al., 2012).

In conclusion, intensive efforts are needed to increase migrants’ knowledge, implement culturally sensitive interventions in migrant communities and better inform health professionals so that they can reach migrants more effectively. Migrants are now a significant part of the population in host countries and appropriate conditions should be created for their best possible adaptation in order to safeguard and promote both migrant and public health.

## Data Availability

All data produced in the present work are contained in the manuscript

